# Participant perceptions of disability training for health workers: a qualitative study in Ghana

**DOI:** 10.1101/2023.11.26.23299018

**Authors:** Sara Rotenberg, Sara Ryan, Sue Ziebland, John Ganle

## Abstract

**Introduction:** Disabled people often report poor treatment by health workers, and health workers often report wanting more training about how to care for disabled people. However, existing disability training for health workers is usually delivered in one-off interventions, with little follow-up, evaluation, and focus on long-term learning. This insufficiency makes it important to understand how disability training for health workers can be more effective. Therefore, we interviewed stakeholders involved in an existing disability training intervention in Ghana to understand how disability training for health workers could be improved.

**Methods:** In-depth, qualitative interviews were conducted with 33 people involved in disability training (either as trainers or trainees) in Ghana. Data were analysed using thematic analysis.

**Results:** Participants spoke about the challenges with existing training, namely how the current approach was insufficient, the consequences of informality in running training and the need for more sign language instruction. Several participants suggested improvements for training, including having external motivation (i.e., professional development credits, monetary benefits, etc.), more collaborative initiatives across institutions and government, and curriculum integration. We developed a theory of change model to show how different components of disability training support learning.

**Discussion:** These results show that disability training for health workers is important and that there is scope to refine and standardize training. In particular, the findings demonstrate how future initiatives to train health workers can be developed and implemented. They also emphasize the need to solicit perspectives from individuals who have experienced training in order to improve future iterations.

## Introduction

Despite the sizable population of disabled people globally, health professional’s curricula are often unsystematic and insufficient with regards to disability training.(1, 2) Accordingly, health workforces often lack adequate knowledge and skills to deliver high-quality health care to disabled people.(3) The omission from training and national policies on sexual and reproductive health (SRH) improvement creates a notable gap in care for disabled women.(4, 5) Ghana, however, has emerged as a leader on research and health worker training on disability and SRH in sub-Saharan Africa, because of training pilots described in this paper.

The 2021 Population and Housing Census of Ghana estimates that 7.8% of the population, or nearly 2.1 million people, are disabled. The most common impairment types are visual (4.0%), physical (3.6%), remembering or concentrating (2.0%) and hearing (1.7%).(6) Disability training for health workers is now included within the Persons with Disability Act, 2006.(7) Despite the commitment to train health workers in disability laws, an implementation gap remains. Indeed, many key health documents have not reflected action on these commitments by ensuring accessibility of outreach and documents, funding, and explicit mentions of disability in policies.(4) The lack of specificity has resulted in an implementation gap in training, as relevant authorities and health worker training schools have not delivered on their obligations under the Ghana Disability Act.(7, 8) Therefore, most training for health workers happens outside of government-focused programs, with academic institutions and Disabled Persons Organisations (DPOs) leading the development and implementation of disability and SRH training for health workers.

### Description of the University of Ghana Training

Since 2017, researchers at the University of Ghana have been involved in developing and delivering disability and SRH training for in-service health workers through various grants. This aimed to improve Ghana Health Service frontline health workers’ understanding of disability and skills to provide family planning and contraceptive services for disabled women and girls. The training was delivered as a two-day course in the Northern District for over 300 health workers in 2017 or 2021.(2) The course introduced disability, disabled peoples’ rights, addressed common misconceptions, and approached SRH topics through a disability lens, including discussing appropriate accommodations for physical examinations, possible contra-indications based on impairments, and appropriate maternal care. Theory-based lectures, role-play and simulations, and disabled guest lectures were the main teaching methods.

The course was initially evaluated with a before and after design and follow-up surveys.(2) This study contributes in-depth interviews with trainees and trainers about the training, its lasting impact, and how this example could improve disability trainings in other contexts. Here we report findings from thematic analysis of these interviews.

## Methods

### Selection and Recruitment of Participants

Interview participants came from two groups: i) trainers and ii) trainees. Trainers included anyone involved in designing and/ or providing disability training for any cadre of health worker. Trainers included disabled people aged 18 years or older from government agencies such as the Ghana Health Service; DPO representatives; health workers; NGOs; and academics. Trainees were from any cadre of health worker (qualified or in-training), whose role maps onto the WHO classification of health workers, who had been involved in the University of Ghana disability training in 2017 or 2021. These two groups represent key stakeholders for health worker training on disability.

Sample size was dictated by saturation of data within the key analytic themes rather than a target sample size.(9) Interviews were conducted with 33 participants in the Greater Accra, Ashanti, and Northern Regions of Ghana. The sample included nine participants with disabilities (Table 1: 27.3%). To capture diverse interview participants, purposeful, snowballing, and convenience sampling methods were used to recruit participants.

**Table 1:** Participant Characteristics.

| N | 33 |
| --- | --- |
| **Gender, n (%)** |  |
| Female | 17 (51.) |
| Male | 16 (48.5) |
| **Place of employment, n (%)** |  |
| Ghana Health Service | 18 (54.5) |
| DPO | 7 (21.2) |
| Academia | 4 (12.1) |
| Hospital | 2 (6.1) |
| Government | 1 (3.0) |
| NGO | 1 (3.0) |
| **Health worker cadre (n=19), n (%)** |  |
| Community Health Nurse | 6 (31.5) |
| Health Volunteer | 4 (21.1) |
| Community Health Officer | 2 (10.5) |
| Midwife | 2 (10.5) |
| Paediatric Nursing Resident | 2 (10.5) |
| Municipal Public Health Nurse | 2 (10.5) |
| District Director | 1 (5.3) |
| **Role in training, n (%)** |  |
| Trainer | 17 (51.5) |
| Trainee | 16 (48.5) |
| **Disability status, n (%)** |  |
| Non-disabled | 24 (72.7) |
| Disabled | 9 (27.3) |

### Data Collection

Semi-structured interviews were conducted in February 2023 in-person, except for one conducted via WhatsApp audio call. Interview questions covered two parts: 1) the training they attended/led and 2) recommendations they would make to improve training. A semi-structured interview guide designed to last 20-30 minutes was used. Interviews were conducted in English with appropriate accessibility measures in place (i.e., sign language interpreter). With consent, interviews were audio recorded and transcribed verbatim.

### Data Analysis

Data were analysed using thematic analysis, (10) which was chosen because it provides a rigorous method for capturing underlying meaning within the data and comparing and contrasting participants’ experiences. Transcripts were checked for accuracy and coded in NVivo, a software to help organize qualitative data. Codes were developed inductively, as well as drawing on the literature and the researcher’s understanding of the topic, to group key topics that recurred in the data. Transcripts were read multiple times to ensure coding was consistent. The OSOP method (10) was used to organise coding extracts to summarise the content of the codes and identify themes to develop inductively. Analysis was led by SR and guided by the overarching question: “what do stakeholders think about the disability training for health workers in which they have been involved?”. The lead author reflected on her personal views, values, and positionality, to challenge any assumptions she was making within the analysis. Final themes were drafted by SR and checked by two authors (SRy and SZ) and rearranged for greater clarity and flow.

### Ethical Considerations

The study was approved by the Oxford Tropical Research Ethics Committee (OxTREC Reference: 534-22) University of Oxford and the Ghana Health Service Ethics Review Committee (Reference: GHS-ERC 005/12/22). Participants were given an information sheet which was read and discussed before participants consented to be interviewed. Data were stored in encrypted formats on secure devices, in-line with ERC requirements.

## Results

Participants reported being pleased with the training they received while suggesting it should come sooner in a health workers’ educational journey. Overall, several ideas for models of good training and suggestions for improvements were expressed. While all aspects of the trainings and the nexus of disability and SRH training were discussed, most participants spoke separately about disability and SRH aspects, making the findings relevant to general disability training.

### Challenges with Training

Participants expressed concerns with the status quo of disability training for health workers in Ghana, including insufficiency, informality, sign language requirements, cost, and lack of coordination to deliver.

### Insufficiency of Current Training Approaches

Most trainers and trainees had been involved in a training for continuing professional development (CPD). However, depending on the health workers’ cadre and school, initial basic disability training varied considerably. For example, a midwife and two nurses demonstrate the variation in training programmes.

> *“In midwifery, it is kind of in passing. You are there to learn things about the gynaecological conditions…they just mention it… [disabled people] are not the majority group in the population so much was not mentioned.”* – Trainee 7, Female, Midwife
>
> *“Yes, it was actually in the nursing education we had. There was a topic on disability – how to identify a disabled person, their weaknesses, and strengths, how to rehabilitate them. But you know because those are always sub-topics, they are not going to really make it so lengthy”* – Trainee 16, Female, Community Health Nurse
>
> *“*Interviewer*: Did you learn about disability in your curriculum?*
>
> Interview Participant*: Actually, no, I didn’t learn.”* – Trainee 14, Female, Community Health Nurse

These quotes highlight the variation that leaves some frontline health workers with inadequate training depending on the institution they attended.

### Consequences of Informality

Several participants highlighted the lack of integration into the curriculum, and difficulty with implementation as sessions were run outside formal roles. Rather, most participants volunteered their time as trainers or had small grants to support the initiative. Trainings were ad-hoc and unsystematic, often run by individuals or DPOs with lived experience, as an informal aspect of the health education system. Most trainers said cost was a substantial challenge particularly because venue, food, lodging, and transportation had to be arranged for participants coming from sub-districts.

> *“I think it is the resources that was making it difficult for us because when you bring them [to Tamale], some are far. They can’t come and go that same day. Because of that we just have to hurry up. We start early so they can go home, but I wish we would have been able to extend the training days”* – Trainer 11, Female, Public Health Nurse

Participants also spoke about the challenge of sustainable funding for training. In one programme participants discussed, training was part of an integrated intervention where participating health volunteers were given motorbikes to help improve disabled peoples’ access to health and transportation to health facilities.(2) However, when the programme ended, there was no funding to continue supporting motorbike costs. This had consequences for health volunteers and disabled people who benefitted from the programme:

> *“Even up to now they are calling, ‘are you people coming to, are you coming again?’ We say, ‘oh no, the project has ended, so maybe unless our boss says we should start again.’ So, they are complaining because some of them, when they want to go to hospital, they will call you ‘Oh can I get help from you to send me to hospital’ so if the program continues it would have been good.” –* Trainee 5, Male, Health Volunteer

Moreover, many trainers noted that the lack of formal recognition from the health education system made it difficult to attract attendees outside of their clinical responsibilities. Some DPOs also noted this lack of integration:

> *“We sent a letter to [hospital] saying we want to come in and offer trainings. They’ve never responded. But I also noticed that when the letters are coming from their top bosses, quickly you get a slot to talk about it. So, can we have this very formalized where we know that ‘ooh okay this is a letter coming from Ministry of Health that [organisation] will be doing a training?’ I think that works, instead of us writing to them directly*” *–* Trainer 5, Female, NGO

While several participants highlighted the difficulty of delivering training on an ad-hoc basis, some felt that even when the training was co-organised with the Ghana Health Service, there was a lack of formal evaluation:

> *“After the training what happens next? Were you able to follow up? Do we really have somebody charged and supported to ensure that the discussion is moved forward? That is a big deficit…so most of the trainings we do—not just on disability—appears to end on the shelf or the training room, even including guidelines that we developed.”* – Trainer 2, Male, Ghana Health Service

Informality of training thus leads to challenges with transportation, costs, recognition, timelines, and evaluation. Trainers said that these challenges affected their ability to reach a broader audience of health workers.

### Sign Language Implementation

Many participants discussed recent efforts to teach sign language to increase communication skills for health workers in Ghana. For some trainees this was their only experience of disability training. However, without formal implementation, the approach was not seen as likely to achieve the goal of expanding communication skills for d/Deaf patients. For example, many clinical participants said that they had learned basic sign language, but without incentives nor opportunities to practice, they forgot the skills learned. This was echoed by a trainer who questioned the time and financial investment involved:

> *“We finished the training for 6 months for mental health nurses and I went to the psychiatric hospital… communicating only in sign language. I could not get a single nurse who could understand my language. So, the question I was asking myself was ‘why did the person come here to spend six months after the lunch, after the water, after the refreshment and coming every week for six good months – 24 weeks – and you still cannot communicate in the basic sign language?’. You are really not helping the institution.” –*Trainer 6, Male, Disabled Persons Organisation

This suggests a need for a more comprehensive approach to maintaining communication skills. One participant was studying sign-language after forgetting the little they had learned in school:

> *“We did sign language in school but not into detail, you know as students we were doing it just to pass our exams and go. But, coming on the field, you realise that you really need the sign language to be able to communicate with them, so I think yes, I am now learning it currently.” –* Trainee 15, Female, Community Health Officer

These extracts demonstrate efforts to improve sign language capabilities while recognizing the challenges of scaling up sign language learning for all health workers, particularly if it is not embedded in practice or examined as part of competency and licensing requirements.

### Suggested Improvements for Training

Participants highlighted aspects of the training that worked well or could be improved. Trainers and trainees said that incentives, collaborative approaches to training, disability-led training, and embedding the training in the health worker curriculums could support increased uptake and efficiency.

### Incentives

Several trainees who had been part of the integrated intervention said that being provided with a motorbike to help disabled people to access services was an effective incentive to participate. They felt it not only helped them better provide services and access clients and facilities, it also served as in important recognition of their work and role in the community. When the study finished it was hard to complete their work without the motorbike and the incentives to visit disabled people in their homes was lost.

Participants also noted that an incentive for using sign language would motivate better language acquisition:

> *“Those with sign language should attract some sort of allowance, you know, additional allowance and it will serve as an inducement.” –* Trainer 16, Female, Government

These tangible incentives were seen as useful tools to improve uptake of health worker training and delivery. In addition, many participants mentioned that more policy-focused incentives would be useful. Some suggested having a training course on disability as part of the licensing requirement or annual CPD requirements for recertification for professional development. Monetary, transport, and certification-focused incentives were seen by participants as being effective tools to improve the uptake of disability training.

### Collaborative approaches

Several trainers had been involved in developing their own curricula and targeting specific organisations to train health workers. They identified two areas where collaborative approaches could strengthen their efforts.

Firstly, many trainers from DPOs said working with the Ghana Health Service or subject-matter experts would lend more credibility and health expertise to their training:

> *“I don’t have a health background so that aspect is given to those with health background to do…the aspect that we bring on board is to let them understand the peculiar nature of disabled people and the fact that they need to be extra careful, and they need to be able to give them the necessary support that they need.” –* Trainer 1, Male, DPO

Similarly, participants from other organisations noted that when disabled people brought their lived experience to the training this provided credibility to the content.

Secondly, for organisations without capacity for learning through patient simulations, role play, etc, a more collaborative approach was seen as important but hard to coordinate without formal partnerships and funding. Through partnering with disabled people learning from classroom-based teaching could be reinforced.

### Train-the-trainer Approaches

Several participants noted that an expectation of doing the training was to share learnings with colleagues, creating an informal community of practice. However, some noted it would be good to have this formalized. Trainees who had previously attended similar training noted that this enabled them to solidify their skills and showed that health workers could help to train their peers:

> *“Those who were trained first became coaches, or let me say facilitators, in the second training. They were now remembering some of the things and teaching those who are just new…they will just come demonstrate and roleplay whatever they learned previously. If there is anything to add, then [the training leaders] show.” –* Trainer 12, Male, Municipal Public Health Nurse

Using a train-the-trainer approach could facilitate regularity and reach of training for health workers. However, neither DPOs nor other training organisations (NGOs, academic institutions, government, etc.) said they could adequately implement training without the other, showing the importance of collaboration to facilitate health worker training.

### Include Disability training in health worker curricula

Finally, many of the health worker participants suggested that while the training helped, disability needed to be included more regularly in their curricula. They hoped this would ensure training reaches all health workers including volunteers:

> *“Every stakeholder involved in health service delivery should have knowledge [on disability] so that at least if the health worker is not there, the volunteer can take charge. [….]in case that person you have trained [..] is not there, otherwise, what happens? The disabled person goes back to the same situation he or she was in.” –* Trainee 5, Female, Community Health Nurse

While there was agreement that there should be curriculum integration, there were different ideas of what would support the best learning. Some said a disability course during training would be sufficient, while others recommended different levels, evaluated for each year of training. Others contended that disability should be central to all training and content, including SRH:

> “*Let them know about the intricacies, you know what [contraceptive] methods they can use what not they cannot use and a little bit about the challenges that might come with that, so that we will be able to give them proper counselling. For instance, somebody will prefer to use a condom, but she doesn’t have dexterity, so she cannot do it alone, but the partner could do that so that you will want to know whether your partner will be willing to play that role you know for you…if we are able to integrate it, into the curriculum like this, then at every point in the curriculum, we look at what will be applicable to people with disability, then we deal with it so that it becomes like we are not doing anything peculiar – it is just part of the normal things that we do.” –* Trainer 14, Male, Academic Physician

The suggestion to integrate specific impairments as part of talking about medical contra-indications creates a new lens with which to integrate and normalise disability in the curriculum. These reflections show various approaches to disability inclusion in health worker education and highlight the need for more systematic integration into the curriculum.

### Theory of Change Model

Participants described being pleased with the approach used to deliver training and elaborated on how methods used were intended to develop (trainers) or did develop (trainees) effective skills. Using deductive and inductive methods, a theory of change model (Table 2) was developed to explore what methods and aspects of training were thought to support acceptable and effective training. The model shows the different elements included in participants’ descriptions of training, validated through comparison with key methods identified in a recent systematic review.(1) The second column highlights the rationale for the element in developing health workers’ skills, largely developed through the analysis of why participants valued the training, and reflections from the literature. The third column shows which health worker outcomes and competencies were intended by the trainers. These results highlight that each component of training has a particular role in building health worker competencies for disability.

**Table 2:** Training Methods Theory of Change Content + Methods – Training.

| **Content + Methods – Training inputs** | **Rationale** | **Intended outcome** |
| --- | --- | --- |
| Review laws and policies | • Establish importance and rights of disabled people.<br>• Set requirement for quality care | • Change behaviour towards disabled people to adhere to laws and deliver rights |
| Exposure to disabled people | • Some health care workers may have never (knowingly) met a disabled person before outside their clinical role.<br>• Cultural beliefs and stigma<br>• Some trainees may have beliefs that disabled people are ‘cursed’ or that disability is ‘contagious’ | • Challenge stigma, attitudes, and beliefs |
| Role play and simulations | • Build confidence in interactions, empathy and understanding.<br>• Practice “bad” and “good interactions in a safe space, with feedback for improvement | • Greater empathy<br>• Better interactions |
| Videos and case studies | • Demonstrate tangible barriers and scenarios for intervention | • Greater understanding |
| Practical skills (i.e., lifts, sighted guiding, etc.) | • Not covered in most health worker training<br>• Provide support in an empowering way, not making assumptions about need for support | • More supportive interactions |
| Sign language and communication skills | • Develop tools to communicate with patients.<br>• Provide greater privacy to disabled people | • Better communication<br>• More privacy |
| Lived experience lectures | • Enhance understanding of disability and barriers to health care<br>• Understand the role of health workers in good-quality health care | • Better understanding of disability and disabled people |
| Manuals and resources | • Support learning outside the classroom<br>• Help learn how to approach similar situations on the go.<br>• Reinforce appropriate language | • Continuous learning on disability |
| Community of Practice | • Build connections between facilities.<br>• Share learnings of ‘what works and positive impact stories | • Refresh skills on disability<br>• Adopt a ‘train-the-trainer’ approach |

## Discussion

This paper discusses perspectives on current disability training, including the challenges, improvements, and methods, that may help lever better training. It is the first study to examine trainers and trainees’ perspectives on disability training for health workers and identifies several aspects of training that could be improved. Overall, there is a dearth of evidence to inform improvements in disability training for health workers, (11) so these findings have implications for how training is designed, implemented, and incentivised.

Overall, these results suggest more attention to disability may be needed in the curriculum. Our data suggest that disability training in Ghana is mainly delivered through ad-hoc initiatives, rather than concerted, systematic, and/or government-led efforts. This is surprising given Ghana has committed to training health workers in its disability laws.(12) While disability training for health workers is not the only gap in implementing these laws, improving integration of these trainings was presented as an important step towards full implementation of the law. Several participants expressed frustration at the sporadic and unequal efforts because of the lack of government leadership in funding disability training within health worker curricula and attaching various policy levers to facilitate its implementation.

However, the suggestion to move towards curriculum integration diverges from what other countries have done to systematically implement health worker training. For example, in the UK, the government has enacted a mandatory program (The Oliver MacGowan Mandatory Training on Learning Disabilities and Autism) for all health and social care staff to take as part of CPD requirements.(13) Similarly, Australia has released a national roadmap for people with intellectual disabilities, which will include developing a required curriculum on intellectual disability for health workers,(14) building on evidence that there has been insufficient inclusion in curricula to date.(15) While many participants in this study said that training was important, some suggested forcing people to do extra training would not yield effective uptake of the skills learned. Instead, integrating it into the curriculum as a lens (like adolescent health or gender) could be more acceptable. This could also help with the sustainability of funding and greater coordination—two substantial barriers to full implementation of the training. Therefore, Ghana and other countries may want to consider these approaches for upskilling health workers, rather than the ad-hoc or mandatory disability-specific CPD courses.

Moreover, our analysis highlights that more policy levers, such as sign language incentives or CPD credits might also facilitate better training, but it is important to note that this may be more appropriate for the current system of ad-hoc training. For instance, CPD credits might be best to incentivise current health workers with no background on disability training to be trained on disability, but that might not be an appropriate incentive once disability is integrated into the curriculum for all health workers. Therefore, more understanding of the impacts of integrating disability into the curriculum is needed and plans for integration should be coupled with appropriate evaluation plans.

Finally, the theory of change model and perspectives on training presented here largely echo the literature surrounding disability competencies in the US(16, 17) and India.(18) That is, many of the suggested methods and curriculum integration would be captured through developing a core set of competencies to which curricula adhered. While there are benefits to developing these competencies at a country-level, such an approach would replicate the unsystematic nature of training at a macro-level, with few similarities between countries. In an era where health workers are more mobile than ever,(19) there is also benefit in developing global standards to ensure similar competencies between all health workers.

### Strengths and Limitations

While most studies have described the immediate impact of training, this study is the first to document trainers and trainees’ views about the successful aspects of training and areas for improvement. By interviewing trainers from a variety of trainings, backgrounds, and geographic locations, we were able to access diverse perspectives on training initiatives across Ghana.

However, since the study was only conducted in Ghana, the findings may not be applicable to every context, particularly where the clinical education system differs. While we tried to recruit a diverse sample, most participants had taken part in the same training which may limit the broader applicability of these findings. It may have been useful to interview some of the non-attendees, as well as observe the training to understand the training fully compared to trainers and trainees accounts.

## Conclusion

These findings show several aspects of existing disability training in Ghana that are relevant to improving future disability training. First, there are several barriers to implementing training, including the insufficiency of existing training, lack of formality, costs and sustainable funding, evaluation, and poor sign language implementation. Potential improvements suggested included: incentives, strong collaboration and coordination, and better curricula integration. In addition, using a combination of teaching methods that have a clear link to the goals of the training will support better training in the future. Ultimately, these results highlight the need to take action to adapt and implement disability training interventions that learn from these barriers, facilitators, and theory of change. By integrating disability training into curricula, health workers will have more opportunities to improve their skills and familiarity with disability and ultimately improve the healthcare experience and outcomes among people with disabilities.

## Data Availability

All data produced in the present study are available upon reasonable request to the authors

